# Haplomics: A Snakemake pipeline for haplotype-based association analysis in multi-omics studies

**DOI:** 10.1101/2025.06.12.25329492

**Authors:** Dariush Ghasemi-Semeskandeh, Luisa Foco, Ryosuke Fujii, Johannes Rainer, Mohadeseh Feli, Markus Ralser, Francisco S. Domingues, Peter P. Pramstaller, Cristian Pattaro

## Abstract

**Summary:** Genome-wide association studies (GWAS) generated thousands of loci associated with complex traits and diseases. However, to characterize of the pleiotropic, molecular and population genetic bounds of uncovered loci, investigations are conducted, that remain conceptually and practically separated. Also in the best case, such investigations proceed by one locus at a time. To address these limitations, we introduce an efficient and reproducible Snakemake pipeline for executing haplotype-based association analysis on GWAS-identified genetic loci, which is especially helpful in samples enriched with molecular omics data. Haplomics takes as input the genetic coordinates of each locus along with all available clinical and molecular phenotypes, the necessary covariates, and VCF genotype files, to reconstruct haplotypes and test them for associations with the phenotypes. The reconstructed haplotypes, the annotation of included variants, and association results are graphically displayed in an HTML report. We tested Haplomics in population-based study sample encompassing 391 traits, including 72 clinical markers, 171 serum metabolites, 148 plasma protein concentrations, and whole-exome sequencing (WES) imputed genotypes. We estimated WES-based haplotypes at 11 kidney function genetic loci from a GWAS and conducted association analyses throughout, identifying 19 significant associations after multiple testing correction. Haplomics is a scalable, easy-to-use and fast haplotype reconstruction and association pipeline that makes it possible to jointly conduct molecular and population-genetic characterization of multiple GWAS loci in unified analysis framework.

**Availability and implementation:** Haplomics is freely available on GitHub at https://github.com/dariushghasemi/haplomics.

## 1. Introduction

Genome-wide association studies (GWAS) identified thousands of loci associated with several complex traits. However, for most identified loci, the relevant biological processes mediating the locus-trait association remain largely uncharacterized [1], requiring subsequent integration of multiple omics (transcriptomics, proteomics, metabolomics) to help identify the most likely causal mechanisms [2]. However, by focusing attention on single-variant analyses, the characterization of the overall variability of identified loci has fallen aside. This includes the evaluation of which and how many genetic profiles at a single locus exist, how they are distributed in population samples, and which phenotypic profiles are associated with the traits of interest. This aspect pertains to the population-genetic aspect of GWAS, which is often seen as separated from the molecular characterization.

Haplotype analysis can help identify the presence of distinct genetic profiles associated with diverse phenotypic manifestations [3], as we also recently showed [4], possibly outperforming single-variant analysis in admixed population samples [5]. Haplotype analysis is rarely considered among post-GWAS investigations as the large number of loci identified by GWAS combined with computationally challenging haplotype reconstruction procedures, generating variable-sized categorical variables, resulting in untreatable computational problems that discourage most researchers.

Workflow management systems such as NextFlow [6], Snakemake [7] and WDL [8] are commonly used to streamline and automate complex analysis workflows. In particular, Snakemake’s Python-based syntax makes it easy to integrate multiple Python tools. Snakemake can perform dry runs, allowing to verify workflows without executing them, ensuring accuracy and avoiding repeated computations. In addition to cluster computing support, Snakemake’s integration with Conda environment and Docker container simplifies dependency management, enhancing reproducibility. Snakemake benefits from extensive documentation and a supportive community, facilitating troubleshooting and learning.

By leveraging on these benefits, we introduce Haplomics, an open-source, efficient Snakemake-based pipeline for conducting haplotype-based association analyses over multiple genetic loci in study samples enriched with multi-omics data. Haplomics unifies genome-scale haplotype analysis with reproducibility. By integrating the R package ‘haplo.stats’ [9], Haplomics enables haplotype reconstruction and subsequent association analysis, allowing a comprehensive exploration of the genetic heterogeneity at all identified loci and their effect on multiple clinical and molecular traits.

Here, we present Haplomics’ design and implementation, illustrating an application to real data in the field of kidney function genetics, where we tested the association between reconstructed whole-exome sequenced (WES)-based haplotypes and multiple clinical and omics phenotypes at 11 loci, with different sizes and characteristics, in a middle-sized population-based study. Eventually, we discuss the gained insights and further extensions.

## 2. Implementation

### Concept outline

The pipeline encompasses a parallelizable series of operations, all based on the combination of five data sources (**Fig. 1a**): (1) individual-level genomic information of all study participants, normally included in a VCF file (typically, this would be the best-guessed imputed genotypes); (2) individual-level clinical and/or omics traits; (3) individual-level covariates (the first necessary genetic principal components (PCs) to control for population structure and any other covariate); (4) map of the genomic regions (loci) within which haplotypes should be reconstructed; and (5) the parameter setting to integrate all provided information.

**Figure 1.**
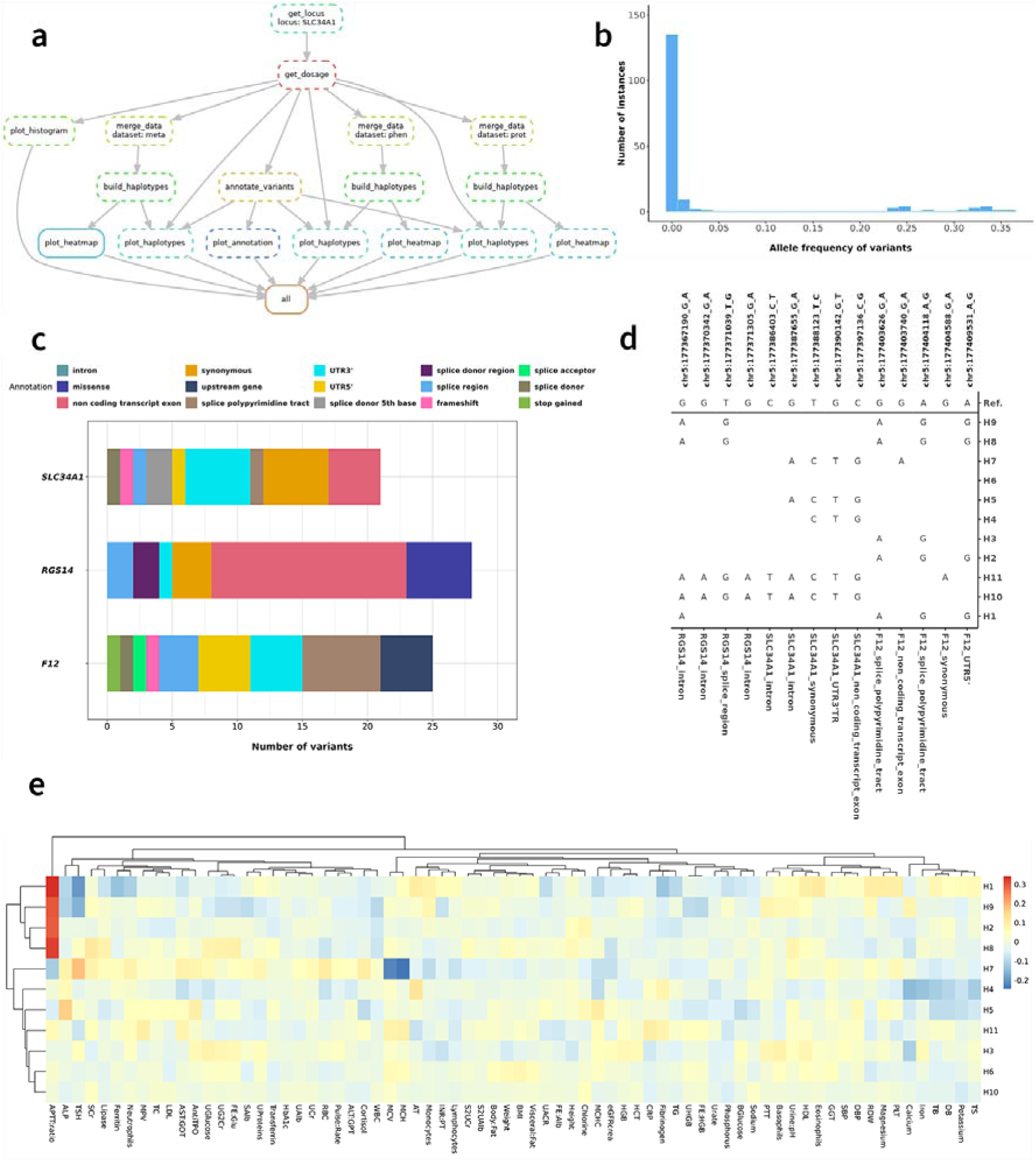
(a) Architecture of the haplotype analysis workflow implemented in Haplomics. (b) Histogram of minor allele frequency of the exonic variants at the *SLC34A1* locus. (c) Bar plot of the most severe consequences of all variants identified for haplotype reconstruction for each gene included in the *SLC34A1* locus. (d) Distribution of the reconstructed haplotypes, identified by tagging variants (multi-allelic variants are removed for clarity) with their functional consequences given on the x-axis. Haplotypes are ordered based on their frequencies reported right of the haplotype label for the analyzed population. (e) Heatmap of z-scores from the associations between haplotypes with any of the analyzed clinical traits.

### Virtual environment

Haplomics installs and loads required packages and command-line tools via Conda, which installs the necessary R packages and bioinformatic tools (**Supplementary Tab. 1**). To allow job parallelization, Snakemake version ≥8.1.1 should be installed. Parallelization is based on the supplied number of cores and exclusive allocation of the required computational resources for each rule of the workflow in a well-formatted configuration file in the slurm/ folder. The pipeline setup and parameter file configuration require basic knowledge of terminal commands.

### Pipeline utilization

The Haplomics pipeline consists of the following 8 steps:

*Step 1. Data extraction.* Chromosomal position, starting and ending genomic coordinates of each locus, and an arbitrary name of the region/locus must be provided in tab-separated file format. Coordinates can be given based on distance, linkage disequilibrium or haplotype blocks [10, 11], recombination hotspots, or other criteria. An indexed subset of the VCF file is generated using *bcftools*.

Step 2. *Annotation*. Best-guessed genotypes, allele frequency, and imputation quality, are extracted for each variant from the subset VCF file via *bcftools*; the most severe consequence of each variant and the respective gene symbol are annotated via the Ensembl Variant Effect Predictor (VEP) API (http://rest.ensembl.org).

Step 3. *Visualization.* The main characteristics of the extracted variants at each locus are displayed via locus-specific histograms of the minor allele frequencies (MAF; **Fig. 1b**) and variant type bar plots by gene (**Fig. 1c Supplementary Fig. S1a-j**).

Step 4. *Data preparation.* The user must provide two data files and their paths through a configuration file in a human readable YAML format: (i) the phenotype file list and (ii) the covariate file (optional). The two files can contain as many traits and covariates as needed. Any appropriate data transformation must be done by the user upfront. To retain equal number of haplotypes for each trait in phenotype file, missing values must be imputed beforehand. To work with multiple datasets with different phenotypes, e.g. from distinct assays, a list of datasets names must be specified in the config file. Haplomics analyzes each assay separately in parallel (unless merging all data upfront).

Step 5. *Association analysis*. Haplotype reconstruction, based on the available variants within each locus, and regression analysis are conducted using the *haplo.glm* function of the R package ‘haplo.stats’ v1.8.9 [9]. Alleles are aligned to the major allele as the reference. Rare haplotype frequency (RHF) must be specified and haplotypes rarer than RHF collapsed into a single rare-haplotype category.

Step 6. *Haplotype plot.* Haplotype variants are listed by chromosomal position, with alleles indicated above the plot and gene name and functional consequences underneath (**Fig. 1d, Supplementary Fig. S2a**). The most common reference haplotype is provided in its entirety on top. For readability, the other haplotypes are presented by listing only the alleles that differ from the reference haplotype.

Step 7. *Association heatmap.* A heatmap is generated, that highlights hierarchical clusters based on the z-scores (effect over its standard error) of the haplotypes-phenotypes associations (**Fig. 1e, Supplementary Fig. S2b-c**).

Step 8. *Result reporting.* For each locus, a user-friendly report is generated that summarizes all generated charts, summary statistics and analysis results. Reports for all loci are integrated into an interactive HTML report, which is documented using a custom R Quarto file and rendered with the rmarkdown v.2.29 R package (**Supplementary File S2-4**). The report gives users the possibility to assess and possibly fine-tune the parameters for analysis improvement. It includes descriptive statistics of the genotype data, tabular association results, visualization of reconstructed haplotypes (full and tagged versions), and a clustered heatmap of the haplotype associations with the available phenotypes in each input dataset. Figures are generated in high-quality PNG format.

### Execution in a high-performance cluster (HPC)

To execute the generated jobs on a HPC computing scheduling system like SLURM, the required settings and parameters are provided in the config.yaml file in the slurm folder.

### Dynamic resource specifications

Step 5 is the most computationally demanding task of the workflow: while a large amount of memory is needed, runtime depends on the locus size. To determine the appropriate resources dynamically, during the workflow execution, Snakemake has a built-in functionality based on the *lambda* or *attempt* expressions. A common example is determining the memory that a job needs, which depends on the input file size of a specific instance. When executing the workflow in a cluster environment, to avoid having the jobs fail due to limited resources, we adopted the built-in parameter *attempt* to adjust resources based on how often the job restarts.

## 3. Results

Haplomics is documented and available at: https://github.com/dariushghasemi/haplomics. To assess its performance, we applied Haplomics to data from the Cooperative Health Research in South Tyrol (CHRIS) study that is a population-based cohort of 13,393 adults aged 18 and over recruited from 13 towns in the alpine Val Venosta/Vinschgau district in the Bolzano-South Tyrol province of northern Italy. In terms of sample size, genomic coverage and number of available traits, the CHRIS study can be considered an average example of population-based studies [12, 13]. In this study, we previously identified 11 genomic loci associated with the estimated glomerular filtration rate (eGFRcrea) [14], replicating results from a large GWAS meta-analysis [15]. In CHRIS, we had 72 available clinical traits and imputed genotypes from a WES panel for 12,834 participants. For 6642 of them, 172 targeted metabolites, quantified in participants’ serum samples, were also available (AbsoluteIDQ p180 kit by Biocrates Life Sciences AG, Innsbruck, Austria). Finally, 3525 participants had available data on 148 plasma protein concentrations (measured using mass-spectrometry-based scanning SWATH [16, 17]). Given the large sample size difference across datasets, we ran three separate analyses: on the clinical phenotypes, on the metabolites, and on the proteins for all 11 loci. All clinical and omics traits were quality controlled and normalized as previously described [4]. Phenotypic missing values were imputed to the median and all phenotypes underwent inverse normal transformation. Variants with minor allele count of <5 were removed and haplotypes with frequency of <0.001 classified as rare. To control the type-I error, we set the significance levels of each analysis as 0.05/(11*72), 0.05/(11*171), and 0.05/(11*148) for clinical traits, metabolites, and proteins, respectively.

The parameter file is provided in **Supplementary File S1**. Mapping of the 11 loci along with their main characteristics is included **Table 1**. Distribution of variants’ MAF for one exemplar locus, *SLC34A1*, are shown in **Fig. 1b** and distribution of variants’ consequences for the SLC34A1 locus in **Fig. 1c**. Functional consequences are presented in **Supplementary Fig. S1a-j**. We fitted linear regression models for each clinical trait, metabolite, and protein, including haplotypes as predictors and adjusting for age, sex, and the first 10 genetic PCs, estimated on the genotyped autosomal variants, to control for population structure. Reconstructed haplotypes for *SLC34A1* are shown in **Fig. 1d** (haplotype tagging variants) and **Supplementary Fig. S2a** (all variants). Associations between haplotypes and phenotypes (**Fig. 1e, Supplementary Fig. S2b-c**). The complete report is uploaded as **Supplementary File S2-4**. The running time distributions and required minimum RAM across the tested loci are described in **Supplementary Tab. S2**. Computational facilities used to conduct the entire analysis by Haplomics are described in **Supplementary Tab. S3.**

**Table 1.**
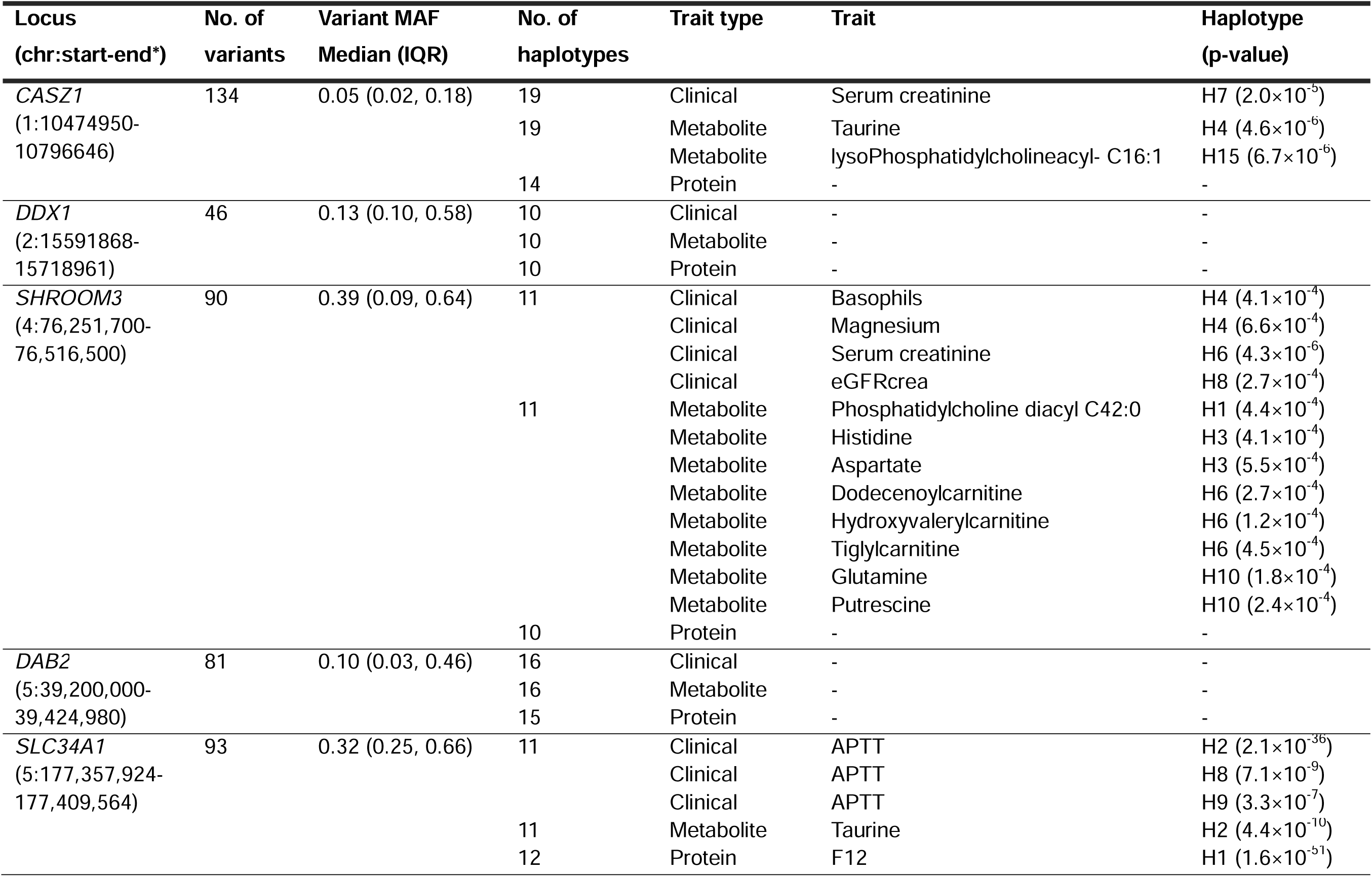
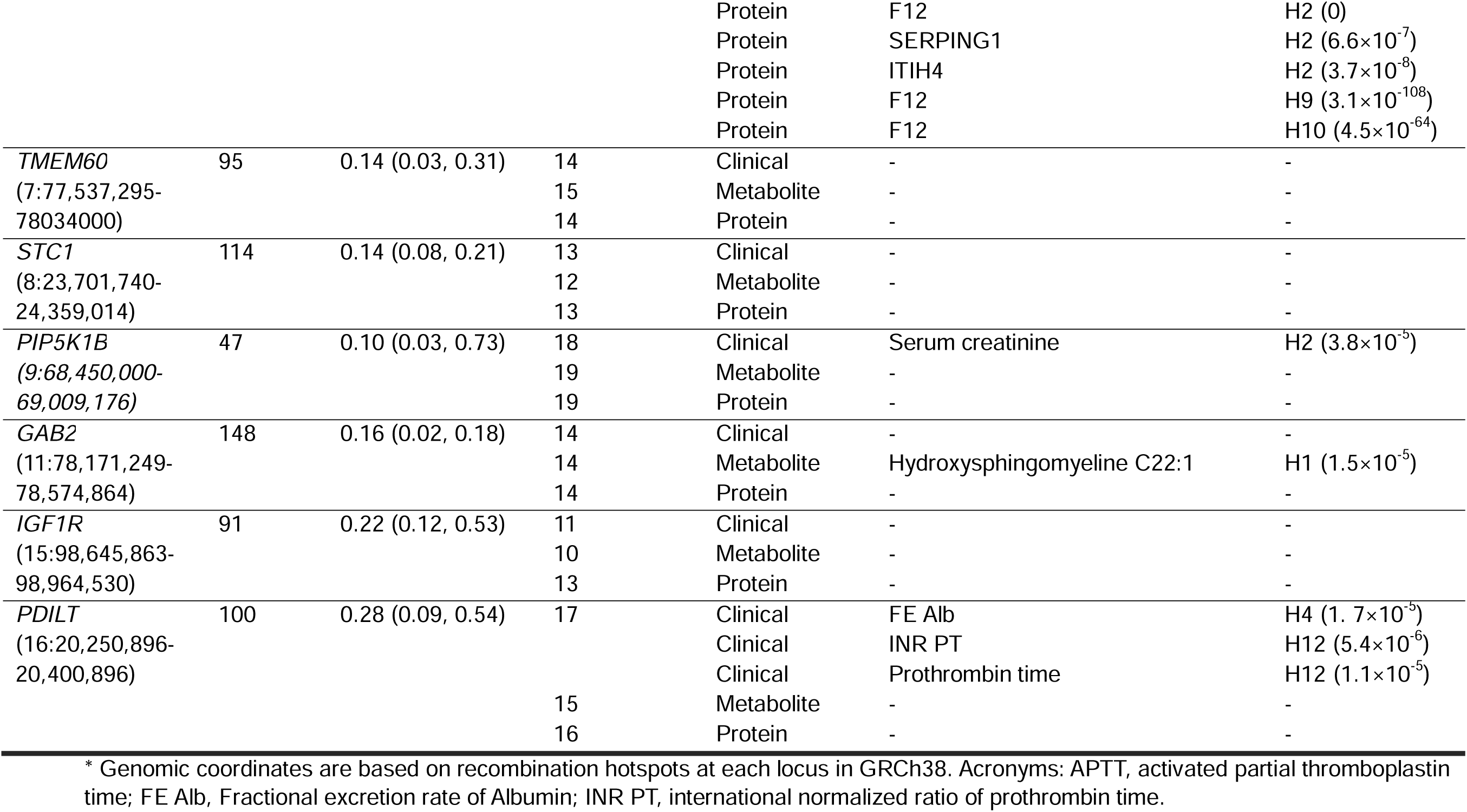
Significant associations between reconstructed haplotypes (allele frequency of >0.01) at the 11 loci for the clinical traits (n=12,834, *P*< 6.2×10^-5^), metabolites (n=6,642, *P*< 2.6×10^-5^) and proteins (n=3,525, *P*< 3.1×10^-5^) in the CHRIS study.

**Table 1** highlights significant associations of haplotypes with clinical traits, metabolites, and proteins. At *SLC34A1*, haplotypes H1, H2, H8, and H9 were associated with higher activated partial thromboplastin time (APTT), connecting to previous research suggesting that genetic variants at this locus might influence kidney function through APTT mechanisms [14]. H1, H2, H9, and H10 were associated with protein levels of coagulation factor *F12* gene at *SLC34A1*, suggesting a potentially shared pathway. H2 was also associated with lower taurine (p<4.39×10^-10^), an endogenous metabolite mainly produced by liver and kidney [18], identified as a potential biomarker of chronic kidney disease (CKD) progression [19]. Overall, Haplomics identified associations providing critical insights into the genetic mechanisms underlying complex physiological relationships (**Supplementary Tab. S4**).

## 4. Discussion

Haplomics operates through a single configuration file with dependencies managed by Conda, enabling straightforward implementation even for researchers with limited bioinformatics expertise. Testing 11 kidney function-associated loci in the CHRIS study revealed 19 significant associations: one the one hand, associations may help elucidate biological mechanisms, on the other hand they highlight the variability of the phenotypic profiles in the general population in relation to the inherited haplotypes [4].

Compared to the very few similar tools, Haplomics offers distinct advantages. CrossHap [20] excels in phylogeny-based visualization while GeneHapR [21] provides visualization features including linkage disequilibrium block analysis and haplotype networks, but both tools lack multi-omics integration. HapTools [5] offers efficient algorithms for population genetic applications. Unlike them, Haplomics provides comprehensive phenotypic association testing across multiple data types and offers more parallelization and high-performance computing integration. Key strengths of Haplomics include its parallelizable workflow design, VCF format compatibility, comprehensive visualization tools, automated resource allocation, and detailed HTML reporting. Limitations are the requirement for terminal command knowledge and dependency on specific R packages that may require updates to maintain compatibility with evolving standards. Additionally, while efficient for small genomic regions and moderate-sized datasets, extremely large cohorts may still require substantial computational resources despite the pipeline’s optimization efforts.

## Supporting information

Supplementary File S2 – Report for clinical traits

Supplementary File S3 – Report for metabolites

Supplementary File S4 – Report for proteins

## Data Availability

The data used in the current study can be requested with an application to: at the Eurac Research Institute for Biomedicine.

https://github.com/dariushghasemi/haplomics

## Acknowledgements

The CHRIS study thanks all study participants, the Healthcare System of the Autonomous Province of Bolzano/Bozen – South Tyrol, and all Eurac Research staff involved in the study (https://www.eurac.edu/chrisack). Bioresource Impact Factor code: BRIF6107. The authors thank the Department of Innovation, Research, University and Museums of the Autonomous Province of Bolzano-South Tyrol for covering the Open Access publication costs.

## Ethics approval and consent to participate

The Ethics Committee of the Healthcare System of the Autonomous Province of Bolzano-South Tyrol approved the CHRIS baseline protocol on 19 April 2011 (21-2011). The study conforms to the Declaration of Helsinki, and with national and institutional legal and ethical requirements. All participants included in the analysis gave written informed consent.

## Supplementary methods

We chose to implement the *haplo.glm* function in the pipeline for several reasons. The haplotype-trait association analysis often involves handling missing phased data, which can be accommodated by the *haplo.glm* reconstruction algorithm. In addition, the generalized linear model (GLM) environment is flexible to incorporate arbitrary numbers of non-genetic covariates and even interactions between genetic and environmental variables. Finally, GLMs can accommodate various types of outcomes including dichotomous, count and continuous traits. Parameter estimates are obtained by use of an expectation-maximization (EM) algorithm for missing categorical covariates [22]. We set the following parameters in *haplo.glm*: n.try=2 (number of times to try to maximize the log-likelihood); insert.batch.size=2 (number of loci to be inserted in a single batch); max.haps.limit=4e6 (maximum number of haplotypes for the input genotypes); and min.posterior=1e-5 (minimum posterior probability for a haplotype pair, given the input genotypes).

**Supplementary Table 1.** Tools and dependencies to run Haplomics on a machine or a cluster environment.

| Tool | Channel | Version |
| --- | --- | --- |
| Snakemake | Conda | ≥8.1.1 |
| Python | Conda | >3.11 |
| requests | Conda | 2.31.0 |
| pandas | Conda | 1.5.3 |
| R | Conda | >3.3.1 |
| Bcftools | Bioconda | 1.21 |
| haplo.stats | R | 1.9.7 |
| pheatmap | R | 1.0.12 |
| tidyverse | R | 2.0.0 |
| data.table | R | 1.15.2 |
| optparse | R | 1.7.4 |
| R.utils | R | 2.12.3 |
| DT | R | 0.32 |
| Digest | R | 0.6.34 |
| rmarkdown | R | 2.29 |
| Quarto | R | 1.12 |
| Knitr | R | 1.45 |

**Supplementary Table 2.** Median (interquartile range) of running time and minimum RAM required for Haplomics by locus and trait groups.

| Locus | Clinical traits |  | Metabolites |  | Proteins |  |
| --- | --- | --- | --- | --- | --- | --- |
|  | Run time (min) | RAM (Gb) | Run time (min) | RAM (Gb) | Run time (min) | RAM (Gb) |
| ***CASZ1*** | 100 (95, 105) | 10 (8, 12) | 111 (106, 116) | 18 (16, 20) | 52 (47, 57) | 10 (8, 12) |
| ***DDX1*** | 40 (35, 45) | 10 (8, 12) | 38 (33, 43) | 18 (16, 20) | 18 (13, 23) | 10 (8, 12) |
| ***SHROOM3*** | 89 (84, 94) | 10 (8, 12) | 62 (57, 67) | 18 (16, 20) | 38 (33, 43) | 10 (8, 12) |
| ***DAB2*** | 77 (72, 82) | 10 (8, 12) | 70 (65, 75) | 18 (16, 20) | 34 (29, 39) | 10 (8, 12) |
| ***SLC34A1*** | 139 (134, 144) | 10 (8, 12) | 78 (73, 83) | 18 (16, 20) | 47 (42, 52) | 10 (8, 12) |
| ***TMEM60*** | 93 (88, 98) | 10 (8, 12) | 76 (71, 81) | 18 (16, 20) | 38 (33, 43) | 10 (8, 12) |
| ***STC1*** | 101 (96, 106) | 10 (8, 12) | 104 (99, 109) | 18 (16, 20) | 51 (46, 56) | 10 (8, 12) |
| ***PIP5K1B*** | 42 (37, 47) | 10 (8, 12) | 55 (50, 60) | 18 (16, 20) | 31 (26, 36) | 10 (8, 12) |
| ***GAB2*** | 116 (111, 121) | 10 (8, 12) | 145 (140, 150) | 18 (16, 20) | 50 (45, 55) | 10 (8, 12) |
| ***IGF1R*** | 109 (103, 113) | 10 (8, 12) | 103 (98, 108) | 18 (16, 20) | 43 (38, 48) | 10 (8, 12) |
| ***PDILT*** | 122 (117, 127) | 10 (8, 12) | 88 (83, 93) | 18 (16, 20) | 47 (42, 52) | 10 (8, 12) |

**Supplementary Table 3.** System configuration of the computing facility employed for the performance evaluation of Haplomics on the study datasets.

| Component | Specification |
| --- | --- |
| Architecture | Linux Infiniband-HDR MIMD Distributed Shared-Memory Cluster |
| Node Interconnect | Infiniband HDR 100 Gb/s |
| Service Network | Ethernet 25 Gb/s |
| CPU Model (CPU nodes) | 2x Intel Xeon Scalable Processors Gold 6226R 2.90 GHz 16 cores |
| Performance (CPU nodes) | 46.58 TFLOPS |
| Operating System | Centos 8.2 - Bright Cluster Manager 9.1 |
| Scheduler | SLURM 20.02.6 |

**Supplementary Table 4.** Bibliographic evidence related to the observed significant associations.

| Locus<br>(putative genes*) | Clinical<br>trait/Metabolite/Protein | Evidence and references |
| --- | --- | --- |
| **CASZ1** (*CASZ1*,<br>*PEX14*) | Serum creatinine<br>Taurine<br>lysoPhosphatidylcholine acyl<br>C16:1 | **Taurine** is an endogenous metabolite mainly produced by liver and kidney (PMID: 31141786), which might be a biomarker of CKD progression (PMID: 30931940). In rats, taurine treatment restored kidney function, reducing blood urea nitrogen (BUN), creatinine levels, and malondialdehyde (MDA), increasing renal levels of superoxide dismutase (SOD), and reversing the increase of kidney injury molecule-1 (KIM-1) and neutrophil gelatinase–associated lipocalin (NGAL) caused by thioacetamide (PMID: 34695366). **LysoPhosphatidylcholine acyl C16:1** (LysoPC C16:1) is a lysophospholipid consisting of one chain of palmitoleic acid at the C-1 position, without known connection with kidney function. The role of the genes in the locus about the identified metabolites and kidney function is unclear. |
| **SHROOM3**<br>(*FAM47E*, *STBD1*,<br>*CCDC158*,<br>*SHROOM3*) | eGFRcrea<br>Serum creatinine<br>Basophils<br>Magnesium<br>Phosphatidylcholine diacyl<br>C42:0<br>Histidine<br>Aspartate<br>Dodecenoylcarnitine<br>Hydroxyvalerylcarnitine<br>Tiglylcarnitine<br>Glutamine<br>Putrescine | The joint associations with clinical traits and metabolites at this locus was extensively discussed in PMID [40485680] |
| **SLC34A1**<br>(*F12*, *GRK6*,<br>*PFN3*, *RGS14*,<br>*SLC34A1*) | APTT<br>Taurine<br>F12<br>SERPING1<br>ITIH4 | **Activated partial thromboplastin time (APTT)** assesses the contact activation and common pathway of blood coagulation cascade. FVIII is inversely associated with eGFR and predicts CKD incident and rapid decline of kidney function (PMID: 30081388). **Taurine** relevance to kidney function is described above. **SERPING1** is an encoded protein of C1-INH, an important regulator of the complement system. SERPING1 peptides may increase in most severe diabetic kidney disease patients' urine (PMID: 31988353) and SERPING1 levels correlate with advanced CKD (PMID: 25589610). **ITIH4** is a liver-produced plasma protein with multiple roles. The best- |
|  |  | known biological function is a protease inhibitor of mannan-binding lectin-associated serine protease-1 (MASP-1), MASP-2, and plasma kallikrein (PMID: 33523981). The plasma peptide of ITIH4 may predict kidney decline in type 1 diabetes patients (PMID: 23466993). |
| **PIP5K1B**<br>(*FAM122A*, *PGM5*,<br>*PIP5K1B*,<br>*TMEM252*) | Serum creatinine | **Serum creatinine** is used to derive eGFR<sub>crea</sub>. *TMEM252* has biased expressions in kidney and other tissues. Common variants near *PIP5K1B* were identified in GWAS of eGFR<sub>crea</sub> and BUN (PMID: 20383146;31152163). |
| **GAB2**<br>(*KCTD21*,<br>*NARS2*, *USP35*,<br>*GAB2*) | Hydroxysphingomyeline<br>C22:1 | **Hydroxysphingomyeline C22:1** is a bioactive sphingolipid abundant in membrane cells and involved in cell growth regulation, cell migration, adhesion, apoptosis, senescence and inflammatory responses (PMID: 38890457). Sphingomyelins were extensively detected in kidney cortex and can be dynamically converted into ceramides, which are linked to acute and chronic kidney disease. The same molecules are involved in multiple physiological kidney function processes (PMID: 39933715). |
| **PDILT**<br>(*UMOD*, *PDILT*,<br>*GP2*) | FE_Alb (Fractional excretion<br>rate of Albumin)<br>INR_PT (international<br>normalized ratio of<br>Prothrombin time)<br>PT (Prothrombin time) | **Fractional excretion rate of albumin** is a known marker of glomerular injury (PMID: 17353515) and a predictor of primary glomerulonephritis progression (PMID: 20921302). **INR_PT** and **PT** measure the activity of the tissue factor (or extrinsic) pathway and the common coagulation pathway. Recent evidence suggests a strong link between coagulation and kidney function (PMID: 26592189). Enrichment in proteins belonging to the canonical pathway of prothrombin activation was found in plasma proteome of CKD patients (PMID: 37816758). More recently, it was observed that the interaction between FXII and urokinase-type plasminogen activator receptor (uPAR) in the kidney tubular cells, activates the integrin β1 pathway, ultimately leading to cell senescence in diabetic kidney disease patients (PMID: 39261453). |

\* Haplotype variants span several genes shown below each locus based on variants annotation using VEP API.

**Supplementary Figure 1.**
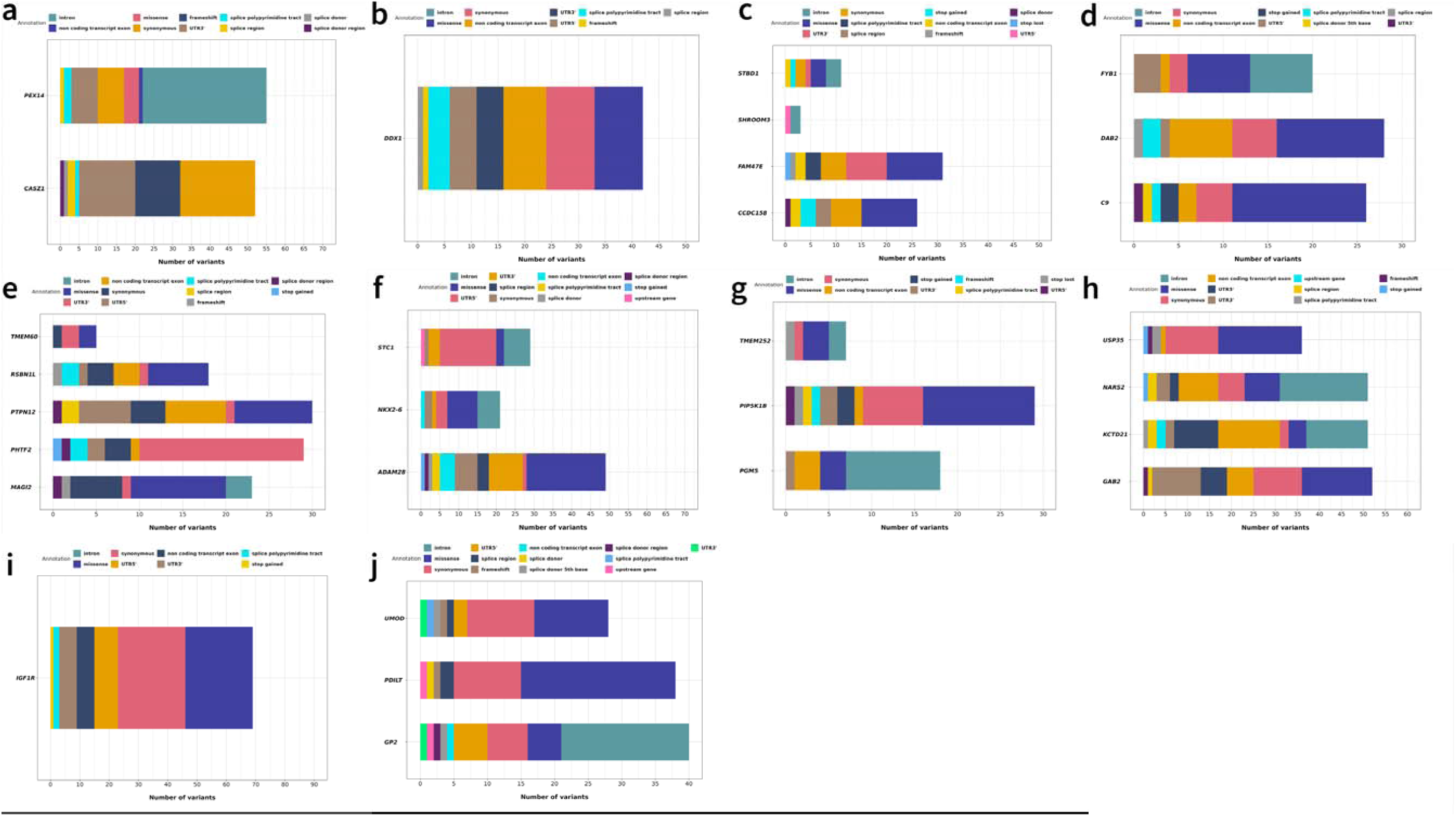
Bar plot of the most severe consequences of all variants identified for haplotype reconstruction for each gene included for each locus in Table 1. (a) *CASZ1* (b) *DDX1* (c) *SHROOM3* (d) *DAB2* (e) *TMEM60* (f) *STC1* (g) *PIP5K1B* (h) *GAB2* (i) *IGF1R* (j) *PDILT*.

**Supplementary Figure 2.**
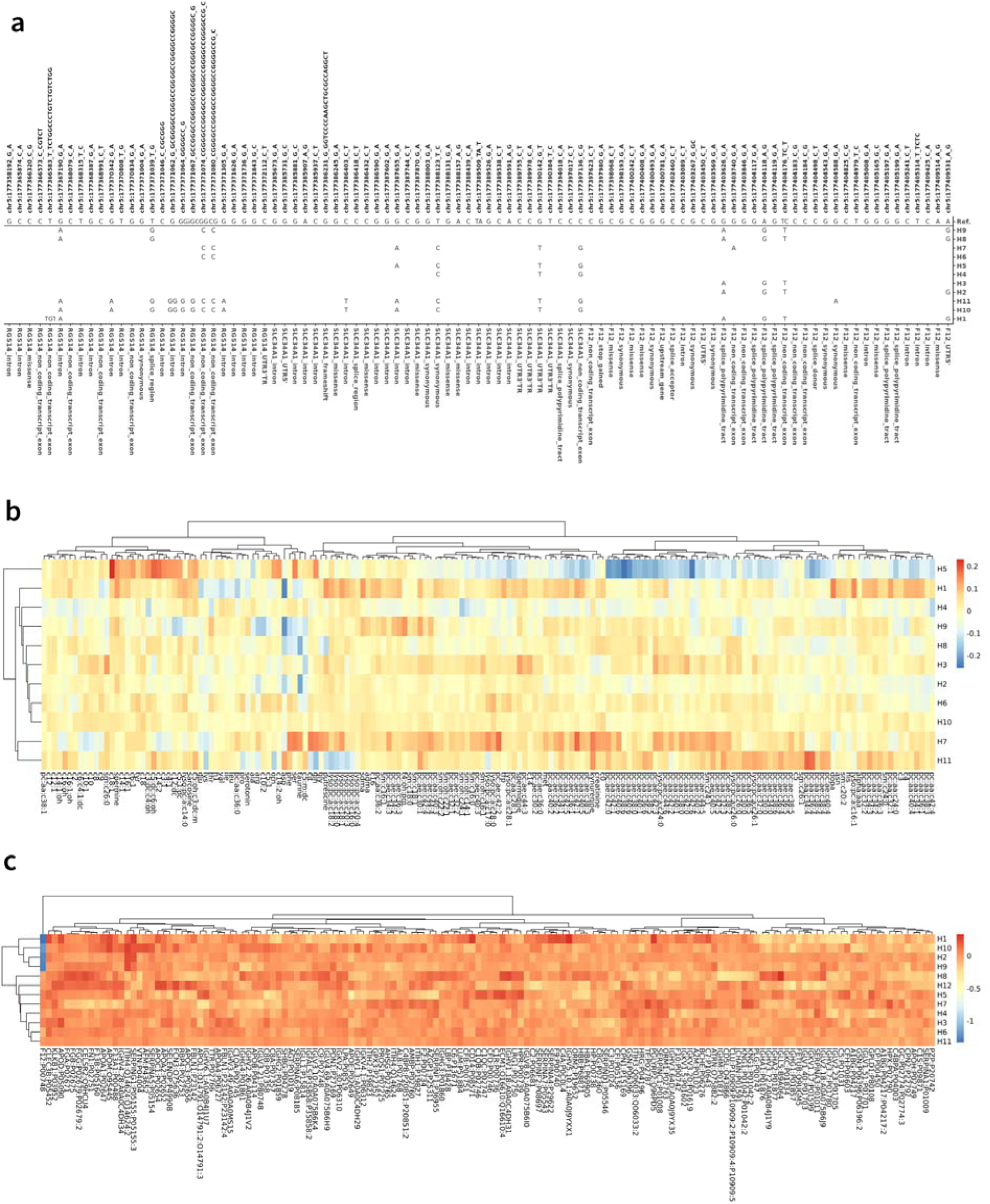
(a) Distribution of the reconstructed haplotypes in their entirety. (b) Heatmap of z-scores from the associations between haplotypes with any of the analyzed serum metabolites. (c) Heatmap of z-scores from the associations between haplotypes with any of the analyzed proteins levels.

**Supplementary File S1.** Structure of the configuration file that user needs to create to run Haplomics on a cluster environment

datasets: “clinical,metabolomics,proteomics”

path_loci: “config/region_coordinates.txt”

path_vcf: “data/WES.imputed.hg38.vcf.gz”

path_workspace: “results/chris”

phenotype_files: “data/clinicals.tsv, data/metabolomics.tsv, data/proteomics.csv”

covariates_file: “data/covariates.tsv”

thresholds:

RHF: 0.001

max_haps: 4e6

min_pp: 1e-5 n_try: 2

n_batch: 2

min_ac: 5

**Supplementary File S2.** HTML report created by Haplomics for *SLC34A1* region with clinical traits dataset

**Supplementary File S3.** HTML report created by Haplomics for *SLC34A1* region with metabolites dataset

**Supplementary File S4.** HTML report created by Haplomics for *SLC34A1* region with proteins dataset

## References

1. Wu, Y., et al., Joint analysis of GWAS and multi-omics QTL summary statistics reveals a large fraction of GWAS signals shared with molecular phenotypes. Cell Genomics, 2023. 3(8).

2. Qi, T., et al., From genetic associations to genes: methods, applications, and challenges. Trends in Genetics, 2024. 40(8): p. 642–667.

3. van der Burg, L.L., et al., Haplotype reconstruction for genetically complex regions with ambiguous genotype calls: Illustration by the KIR gene region. Genetic Epidemiology, 2024. 48(1): p. 3–26.

4. Ghasemi-Semeskandeh, D., et al., Clinical and Metabolic Signatures of FAM47E-SHROOM3 Haplotypes in a General Population Sample. Kidney International Reports, 2025. 10(5): p. 1495–1508.

5. Massarat, A.R., et al., Haptools: a toolkit for admixture and haplotype analysis. Bioinformatics, 2023. 39(3).

6. Di Tommaso, P., et al., Nextflow enables reproducible computational workflows. Nature biotechnology, 2017. 35(4): p. 316–319.

7. Köster, J. and S. Rahmann, Snakemake—a scalable bioinformatics workflow engine. Bioinformatics, 2012. 28(19): p. 2520–2522.

8. Voss, K., J. Gentry, and G. Van Der Auwera, Full-stack genomics pipelining with GATK4+ WDL+ Cromwell. F1000Research, 2017. 6(1381).

9. Schaid, D.J., et al., Score Tests for Association between Traits and Haplotypes when Linkage Phase Is Ambiguous. The American Journal of Human Genetics, 2002. 70(2): p. 425–434.

10. Pattaro, C., et al., Haplotype block partitioning as a tool for dimensionality reduction in SNP association studies. BMC Genomics, 2008. 9: p. 405.

11. Taliun, D., J. Gamper, and C. Pattaro, Efficient haplotype block recognition of very long and dense genetic sequences. BMC Bioinformatics, 2014. 15: p. 10.

12. Pattaro, C., et al., The Cooperative Health Research in South Tyrol (CHRIS) study: rationale, objectives, and preliminary results. Journal of Translational Medicine, 2015. 13(1): p. 348.

13. Lundin, R., et al., Cohort Profile: the Cooperative Health Research in South Tyrol study. Int J Epidemiol, 2025. 54(3).

14. Ghasemi-Semeskandeh, D., et al., Systematic mediation and interaction analyses in an individual population study help characterize kidney function genetic loci. medRxiv, 2023: p. 2023.04.15.23288540.

15. Wuttke, M., et al., A catalog of genetic loci associated with kidney function from analyses of a million individuals. Nat Genet, 2019. 51(6): p. 957–972.

16. Messner, C.B., et al., Ultra-fast proteomics with Scanning SWATH. Nat Biotechnol, 2021. 39(7): p. 846–854.

17. Dordevic, N., et al., Pervasive Influence of Hormonal Contraceptives on the Human Plasma Proteome in a Broad Population Study. medRxiv, 2023: p. 2023.10.11.23296871.

18. Jakaria, M., et al., Taurine and its analogs in neurological disorders: Focus on therapeutic potential and molecular mechanisms. Redox Biol, 2019. 24: p. 101223.

19. Chen, D.Q., et al., Identification of serum metabolites associating with chronic kidney disease progression and anti-fibrotic effect of 5-methoxytryptophan. Nat Commun, 2019. 10(1): p. 1476.

20. Marsh, J.I., et al., *crosshap: R package for local haplotype visualization for trait association analysis*. Bioinformatics, 2023. 39(8).

21. Zhang, R., G. Jia, and X. Diao, geneHapR: an R package for gene haplotypic statistics and visualization. BMC Bioinformatics, 2023. 24(1): p. 199.

22. Burkett, K., J. Graham, and B. McNeney, *hapassoc: Software for likelihood inference of trait associations with SNP haplotypes and other attributes*. Journal of Statistical Software, 2006. 16: p. 1–19.

