## Supplementary File S2 – Report for clinical traits for "Haplomics: A Snakemake pipeline for haplotype-based association analysis in multi-omics studies"

Haplomics report for clinical at SLC34A1.


Code 

- Show All Code
- Hide All Code
- Download Rmd

### Haplomics report for clinical at *SLC34A1*.

This report is generated by Quarto on June 10, 2025


##### Introduction

This report demonstrates results of the conducted analysis for
clinical at *SLC34A1*.

##### Minor allele frequency of variants


Figure 1. Allele frequency of the variants

##### Variants annotation plot


Figure 2. Number and annotation of the variants in each gene


###### Discriptive statistics


Minimum allele count (AC) is set to 5 . Given 13447 samples, variants
with AC below 5 and above 13442 are removed., , , Of 165 variants, 93
left with minor allele count > 5 for building haplotypes., , , Merged
data comprised 93 variants, 73 traits, and 13 covariates.,

##### Haplotype plot

###### Haplotype variants


Figure 3. Alleles distribution across haplotypes

###### Haplotype tagging variants


Figure 4. Haplotypes narrowed by varied variants

##### Associations with clinical

###### Heatmap plot


Figure 5. Heatmap plot of the associations coefficients

###### Table of results
